# Objective quantitation of EGFR protein levels using Quantitative Dot Blot (QDB) method for prognosis of gastric cancer patients

**DOI:** 10.1101/2021.04.12.21255321

**Authors:** Lei Xin, Fangrong Tang, Bo Song, Maozhou Yang, Jiandi Zhang

**Author notes:** Correspondence should be address to at Quanticision, Diagnostics, Inc. of RTP, NC, USA. These authors contributed equally (Lei Xin, Fangrong Tang).

## Abstract

**Background:** One causing factor underlying failures of several clinical trials of anti-EGFR therapies is the lack of effective method to select patients overexpressing EGFR protein. Quantitative Dot Blot method (QDB) is proposed here to measure EGFR protein levels objectively and quantitatively. Its feasibility was evaluated for prognosis of overall survival (OS) of gastric cancer patients.

**Methods:** Formalin Fixed Paraffin Embedded (FFPE) slices of 2×5 μm from gastric and Lung cancer specimens were used to extract total tissue lysate for QDB measurement. Absolutely quantitated EGFR protein levels were used for Kaplan-Meier Overall Survival (OS) analysis of gastric cancer patients.

**Results:** EGFR protein levels ranged from 0 to 772 pmole/g (n=246) for gastric, and from 0 to 2695 pmole/g (n=81) for lung cancer patients. Poor correlation was observed between quantitated EGFR levels and immunohistochemistry (IHC) scores with r=0.018, p=0.786 from Spearman’s correlation analysis. EGFR was identified as an independent negative prognostic biomarker for gastric patients only through absolute quantitation, with HR at 2.29 (95%CI:1.23-4.26, p=0.0089) from multivariate cox regression OS analysis. A cutoff of 207.7 pmole/g was proposed to stratify gastric cancer patients, with 5-year survival probability at 37% for those whose EGFR levels were above the cutoff, and at 64% those below the cutoff based on Kaplan-Meier OS analysis. p=0.0057 from Log Rank test.

**Conclusion:** A QDB-based assay was developed for both gastric and Lung cancer specimens to measure EGFR protein levels absolutely, quantitatively and objectively. This assay should facilitate clinical trials aiming to evaluate anti-EGFR therapies retrospectively and prospectively.

## Introduction

Aberrant overexpression of Human epidermal growth factor receptor (EGFR) protein has been documented extensively in tumorigenesis, growth and progression in various types of cancers (1–3). Anti-EGFR monoclonal antibody drugs, including Cetuximab, Panitumumab, Nimotuzumab and Necitumumab, have been developed to treat cancer patients in daily clinical practice (4).

Conceivably, the efficacy of all these monoclonal antibody drugs should be tightly associated with EGFR protein levels at tumor tissues, considering EGFR is the antigen of these antibodies. As expected, there were evidences suggesting these anti-EGFR monoclonal antibody drugs performed better among patients overexpressing EGFR proteins than those with low to moderate levels (2,5–9). However, there are also a large number of studies suggesting EGFR was not an effective predictive biomarker for these monoclonal antibody drugs (7,10–14). In several clinical trials, these monoclonal antibody drugs were administered to participants regardless of their EGFR protein levels (2,11,15–17).

It is rather counter-intuitive to think that a monoclonal antibody drug may be working in the absence of its antigen. While we may attribute this unexplainable phenomenon to the complexity of cancer biology, the possibility of faulty detection method cannot be dismissed easily.

Currently, the EGFR protein levels are assessed primarily with immunochemistry (IHC), a semi-quantitative method known to be associated with subjectivity and inconsistency(2,18). In majority of the studies, the results of IHC analysis, or IHC scores, of EGFR protein expression are categorized as 0/1+/2+/3+ based on the number of positively stained cells and the intensities of staining. A more complicated method (H score) is also used in several retrospective clinical studies by multiplying the intensity (from 0 to 3) of the signal with the corresponding portion of the cells (from 0 to 100%) to yield a grading score ranging from 0 to 300 (5–8,10). However, this practice may introduce more subjectivity in these studies.

The other issue with IHC method is its ineffectiveness to define the right threshold to separate EGFR positive (EGFR+) from EGFR negative (EGFR-) patients due to its oversimplified categorization strategy (12). In most cases, patients with any expression of EGFR protein are defined as EGFR+ (IHC score ≥1+). In other studies, the cut-offs were set around 200, 220 and 240 in the 0∼ 300 scale based on limited outcome analyses (5,7–9).

We hypothesized that the subjective and semi-quantitative nature of IHC method at least partly undermine the efforts to extend these anti-EGFR therapies to treat gastric and Non-small Cell Lung Cancer (NSCLC) patients (2,7,8). Clearly, novel method offering objective quantitation of EGFR protein levels is needed to address these issues. The quantitated results will also help us to develop the optimized cutoff objectively through outcome analysis to guide patients with these anti-EGFR therapies.

Recently, Quantitative Dot Blot method (QDB) has been used to measure Her2 and Ki67 protein levels absolutely and quantitatively in breast cancer specimens. This is an ELISA-like method ready to be adopted in routine clinical practice(19–22). Compared with IHC method, QDB method is more simple, objective and consistent. It also provides absolute quantitated protein levels for unmatched accuracy over IHC method.

In this study, we introduced a QDB-based assay to measure EGFR protein levels absolutely and quantitatively in gastric and lung cancer tissues. The feasibility of this method was evaluated by exploring EGFR as a prognostic biomarker for gastric cancer patients through overall survival (OS) analysis based on its absolutely quantitated protein levels.

## Materials & Methods

### Human subjects

A total of 248 Formalin Fixed Paraffin Embedded (FFPE) gastric specimens and 81 FFPE lung cancer specimens as 2×5µm slices between Jan. 2015 and Dec. 2017 were provided sequentially and non-selectively by Yantaishan Hospital at Yantai, P. R, China. All the gastric cancer tissues were from patients at M_0_ stage, and the flow chart was shown in Fig. 1. The study was conducted in accordance with the Declaration of Helsinki, following a protocol approved by ethics committee of Yantaishan hospital (YanshanLunZhun2021017 to Lei Xin), with an informed consent waiver due to the use of archival tissues with retrospective, anonymized clinical data.

**Fig. 1:**
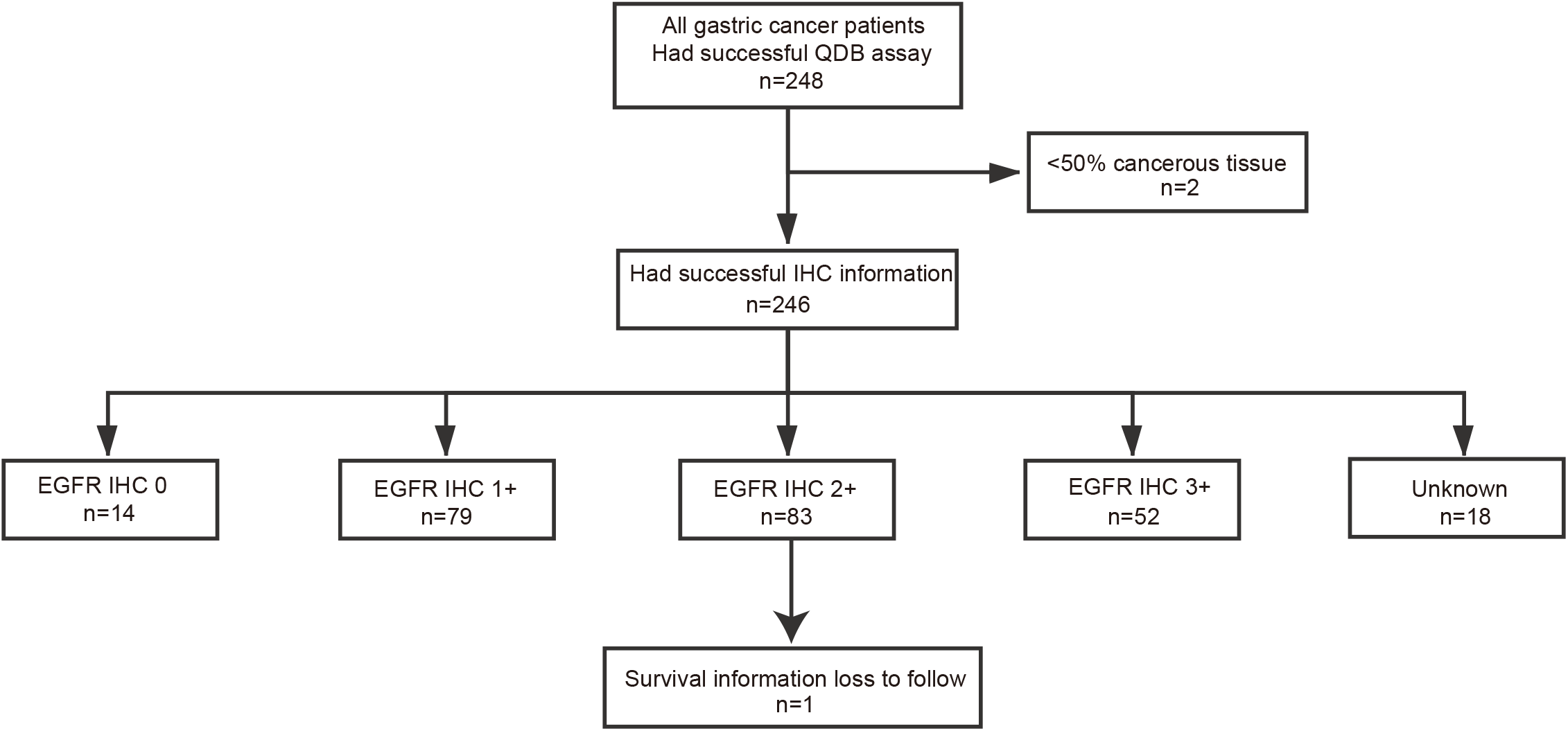
Flow chart of the gastric cancer specimens

### General reagents

All of the chemicals were purchased from Sinopharm Chemicals (Beijing, P. R. China). Recombinant human EGFR protein was purchased from Sino Biological Inc. (Cat#: 10001-H20B2, Beijing, China). Rabbit anti-EGFR antibody (clone EP22) was purchased from ZSGB-BIO (Beijing, China). HRP labeled Donkey Anti-Rabbit IgG secondary antibody was purchased from Jackson Immunoresearch lab (West Grove, PA, USA). BCA protein quantification kit was purchased from Thermo Fisher Scientific Inc (Calsband, CA, USA). QDB plate was provided by Quanticision Diagnostics, Inc (RTP, USA).

### Preparation of FFPE tissue

Two FFPE tissue slices at 5µm each (2×5µm) were collected into 1.5ml Eppendorf tubes, and de-paraffinized before being solubilized using lysis buffer (50mM HEPES, 137mM NaCl, 5mM EDTA, 1mM MgCl_2_, 10mM Na_2_P_2_O_7_, 1% TritonX-100, 10% glycerol). FFPE tissue slices of a gastrointestinal stromal tumor (GIST) were also lysed in the same lysis buffer to serve as the internal control throughout the study. Supernatants were collected after centrifugation and the total amount of proteins was determined using BCA protein assay kit.

### Developing QDB-based EGFR assay

Total FFPE tissue lysates from 60 ∼ 80 gastric and lung cancer specimens respectively at 0.5μg/unit were loaded onto QDB plate. The quality of the detection antibody (EP22 at 1:1000 dilution) was evaluated first by screening the signals of these specimens over the blank control (Bovine serum albumin at 0.5μg/unit). After screening, total tissue lysates from 4 to 6 specimens of middle range expression were pooled together, and used to develop a dose curve, alongside with the serial diluted purified EGFR proteins, as shown in supplemental fig.1a, 1b & 1c. The optimized loading amount of the total protein was used for absolute quantitation of EGFR protein levels in gastric or lung cancer tissues.

### QDB analysis

The QDB process was described elsewhere with minor modifications(21). In brief, the final concentration of the FFPE tissue lysates was adjusted to 0.25 µg/µl, and 2 µl/unit was used for QDB analysis in triplicate. The loaded QDB plate was dried at room temperature (RT) for an hour before it was blocked in blocking buffer (4% non-fat milk in TBST) for an hour. The plate was inserted into a 96-well microplate filled with 100µl/well primary antibody (for clone EP22, 1:1000 in blocking buffer), and incubated overnight at 4^0^C. Afterward, the plate was rinsed twice with TBST and washed 3×10 minutes before it was incubated with a donkey anti-rabbit secondary antibody for 3 hours at RT. The plate was rinsed twice with TBST, washed 5×10 minutes and then was inserted into a white 96-well plate pre-filled with 100µl/well ECL working solution for 3 minutes. The chemiluminescent signals of the combined plate were quantified using Tecan Infinite 200pro Microplate reader with the option “plate with cover”. The results were average of three independent experiments.

The GIST lysate with pre-documented EGFR level was included as internal control in all the assays to ensure the consistency of the results. The individual assay was accepted only when measured EGFR level of GIST internal control was within 80%∼120% of pre-determined level. Samples with chemiluminescent reading less than 2 fold over blank were defined as non-detectable, and entered as 0 for data analysis.

### Statistical analysis

All statistics were performed using GraphPad Prism software version 7.0 (GraphPad Software Inc., USA) and R version 4.0.4, using two-side statistical test. Missing values in discrete data were defined as a new category. The results were reported as mean ± standard Deviation (SD). p values of less than 0.05 were considered statistically significant. The endpoint of overall survival (OS) analysis was defined as the time from surgery to death or the last follow-up. The last follow-up data was Nov.11st, 2020. Patients who lost to follow-up were excluded. Survival data for patients who were still alive at the date of last follow-up were treated as censored.

Univariate Cox proportional hazard models fitted overall survival were employed for hazard ratio (HR) and corresponding 95% confidence intervals (CIs) estimation. Multivariable Cox models were utilized to examine the association between subtypes and OS, adjusting for other clinical variables, such as age, node status, tumor size, tumor grade, and type of treatment. Residuals that are analogous to the Schoenfeld residuals in Cox models were used to check the proportionality assumption.

## Results

### Measuring EGFR protein levels in gastric and lung cancer specimens

Clinicopathological characteristics of all gastric and lung cancer specimens were listed in Table 1 and supplemental table 1. The optimization of QDB-based EGFR assay was detailed in Materials and Methods section. Two 5 μm (2×5μm) FFPE slices with more than 50% tumor tissues (based on H & E staining) from individual gastric or Lung cancer specimen were used for total protein extraction. The EGFR protein level in each specimen was measured as detailed in QDB assay using clone EP22 as the primary detection antibody. The distributions of EGFR protein levels in gastric cancer specimens (n=246) and Lung cancer specimens (n=81) were shown in Fig. **2a**.

**Table 1:** Clinicopathological characteristics of gastric cancer specimens.

| **Characteristics** |  | **Cases (n=246)** |  |
| --- | --- | --- | --- |
|  |  | **n** | **%** |
| **Age** |  |  |  |
|  | <60 | 88 | 35.77 |
|  | ≥60 | 158 | 64.23 |
| **Gender** |  |  |  |
|  | Male | 180 | 73.17 |
|  | Female | 66 | 26.83 |
| **Histologic grade** |  |  |  |
|  | well/moderately differentiated | 25 | 10.16 |
|  | poorly differentiated | 221 | 89.84 |
| **Tumor size** |  |  |  |
|  | Small(<5cm) | 120 | 48.78 |
|  | Large(≥5cm) | 120 | 48.78 |
|  | Unknown | 6 | 2.44 |
| **Lymph node metastasis** |  |  |  |
|  | No metastasis | 88 | 35.77 |
|  | Metastasis | 158 | 64.23 |
| **T-stage** |  |  |  |
|  | 1 | 50 | 20.33 |
|  | 2 | 22 | 8.94 |
|  | 3 | 54 | 21.95 |
|  | 4 | 120 | 48.78 |
| **LNsn** |  |  |  |
|  | N0 | 88 | 35.77 |
|  | N1 | 39 | 15.85 |
|  | N2 | 38 | 15.45 |
|  | N3 | 81 | 32.93 |
| **Vascular cancer embolus** |  |  |  |
|  | Yes | 123 | 50.00 |
|  | No | 119 | 48.37 |
|  | Unknown | 4 | 1.63 |
| **Nerve invasion** |  |  |  |
|  | Yes | 102 | 41.46 |
|  | No | 140 | 56.91 |
|  | Unknown | 4 | 1.63 |
| **Lauren's classification** |  |  |  |
|  | intestinal type | 50 | 20.33 |
|  | diffuse type | 85 | 34.55 |
|  | mixed type | 84 | 34.15 |
|  | Unknown | 27 | 10.98 |
| **EGFR IHC** |  |  |  |
|  | 0 | 14 | 5.69 |
|  | 1+ | 79 | 32.11 |
|  | 2+ | 83 | 33.74 |
|  | 3+ | 52 | 21.14 |
|  | Unknown | 18 | 7.32 |
| **Survival** |  |  |  |
|  | Yes | 156 | 63.41 |
|  | No | 89 | 36.18 |
|  | Unknown | 1 | 0.41 |

**Fig. 2.**
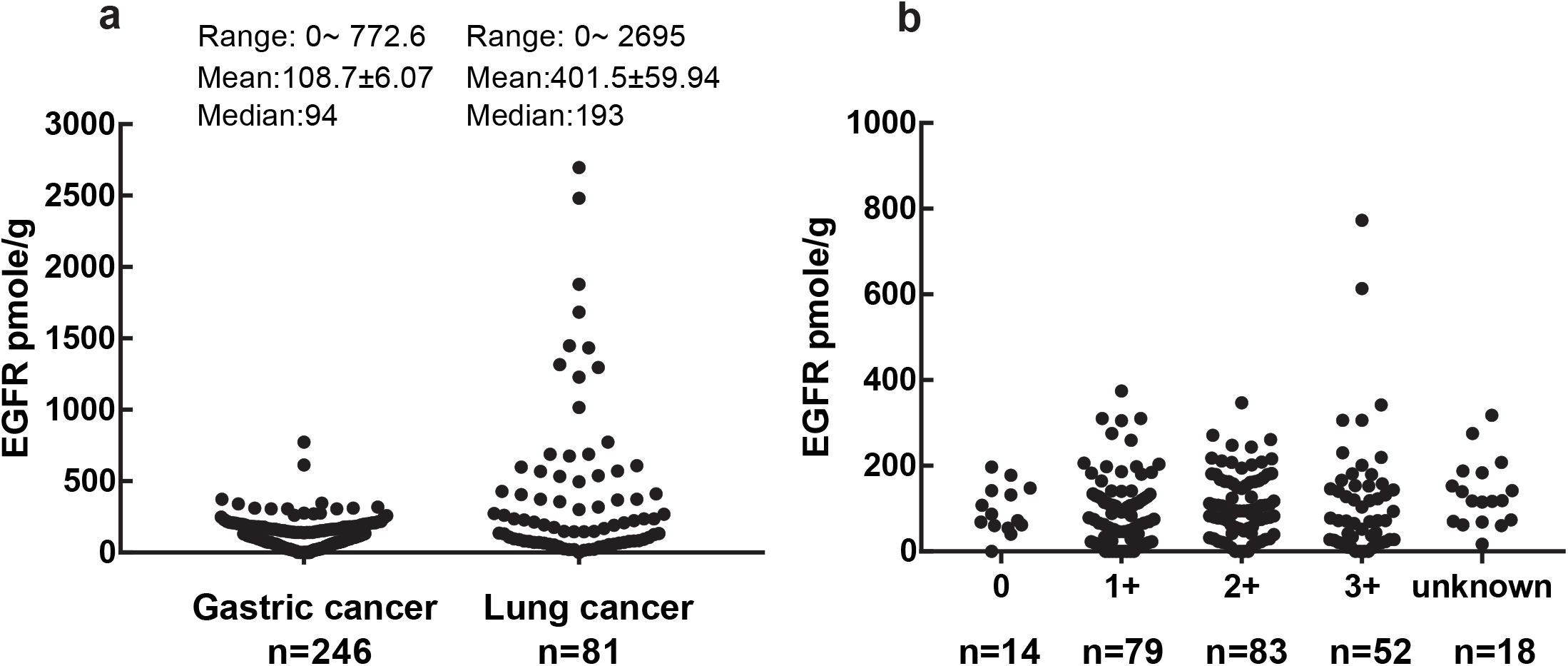
Distribution of absolutely quantitated EGFR protein levels in gastric and lung cancer specimens (**a**), and their distribution grouped by their IHC scores among gastric cancer specimens (**b**). Total protein lysates were prepared from 2×5 µm FFPE slices for QDB measurement using Rabbit monoclonal anti-EGFR antibody (EP22). In Fig 1b, EGFR protein levels of individual FFPE gastric cancer specimens were grouped based on their IHC scores. There was no statistical significance between any two IHC groups when analyzed using Student t-test.

Among gastric cancer specimens (n=246), the EGFR protein levels ranged from 0 (undetectable level) to 772.6 pmole/g, with median at 94 pmole/g, and mean at 108.7± 6.07 pmole/g. For Lung cancer specimens, the EGFR protein levels ranged from 0 to 2695 pmole/g, with median at 193 pmole/g, and mean at 401.5±59.94 pmole/g.

The absolute quantitated EGFR levels were also plotted against their respective IHC scores in Fig. **2b**. In 0 group (n=14), EGFR levels were from 0 to 197 pmole/g, with average at 96.54 pmole/g; in 1+ group (n=79), EGFR levels were from 0 to 374.4 pmole/g, with mean at 97.9 pmole/g; in 2+ group (n=83), EGFR levels were from 0 to 347.5 pmole/g, with mean at 109 pmole/g; and at 3+ group (n=52), EGFR levels were from 0 to 772.6 pmole/g, with mean at 118.9 pmole/g. No statistical difference was detected between any two IHC groups when analyzed with student t-test (data not shown). When analyzed with Spearman’s correlation analysis, the results from QDB method were associated poorly with IHC scores, with r=0.018, p=0.786.

Next, we investigated the prognostic role of EGFR in gastric cancer through both univariate and multivariate OS analysis (table 2). The EGFR protein levels were negatively associated with OS, with HR at 2.15 (95%CI: 1.23-3.75, p=0.0069) for univariate, and 2.29 (95%CI: 1.23-4.26, p=0.0089) for multivariate cox regression OS analysis respectively. In addition, lymph node status was also found to be an independent prognostic factor in multivariate cox regression OS analysis. It should be mentioned that IHC scores were found not an independent prognostic factor in neither univariate nor multivariate cox regression OS analysis (Supplemental table 2).

**Table 2:**
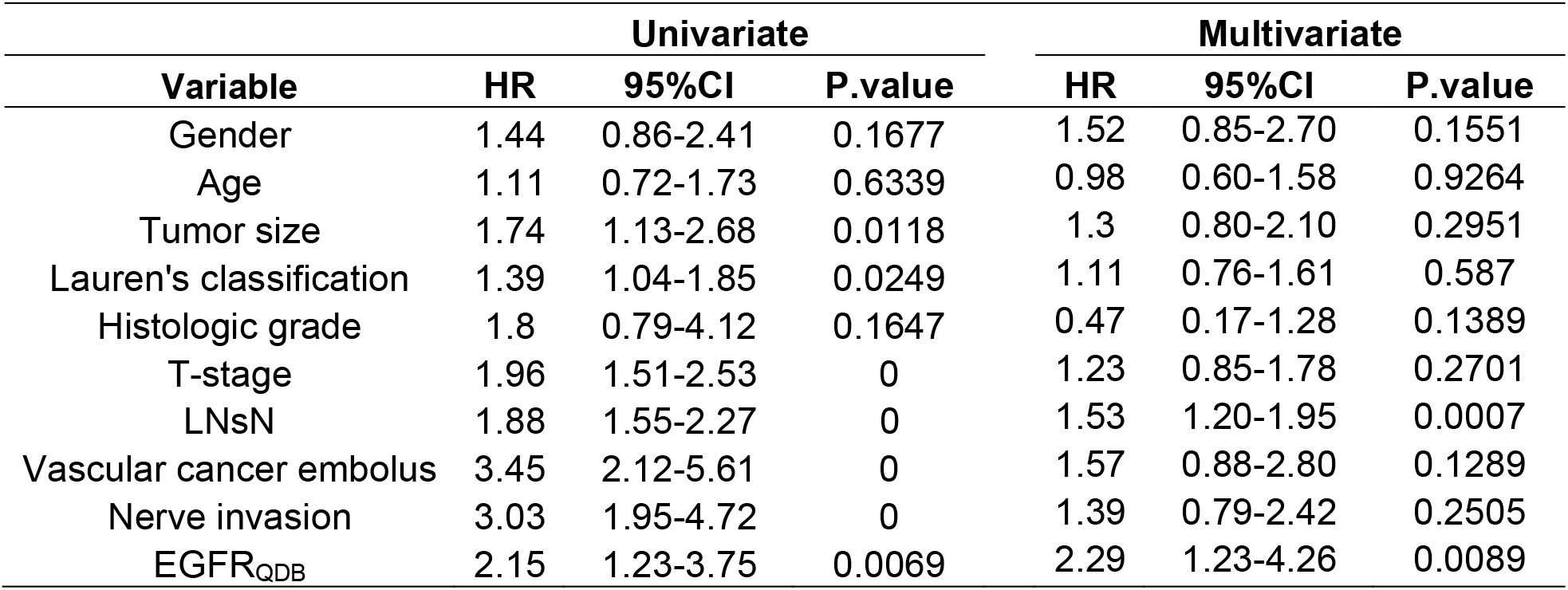
Univariate and Multivariate Cox regression of Overall Survival (OS) with EGFR levels.

Having demonstrated EGFR as an independent prognostic factor for gastric cancer patients, a putative cutoff was explored to predict 5 year survival probability (5y SP) between patients with their EGFR levels above the cutoff (EGFR+) and those below cutoff (EGFR-). The “surv_cutpoint” function of the “suvminer” R package in combination with the OS of these patients was used for this search to identify a putative cutoff of 207.7 pmole/g. In Fig. 3, we evaluated the effectiveness of this putative cutoff using Kaplan-Meier analysis. As shown in Fig. **3a**, EGFR-patients had 5y SP at 64% (95%CI: 57% to 72%), in contrast to EGFR+ patients with 5y SP at 37% (95%CI: 21% to 65%). p=0.0057 from Log Rank test.

**Fig. 3:**
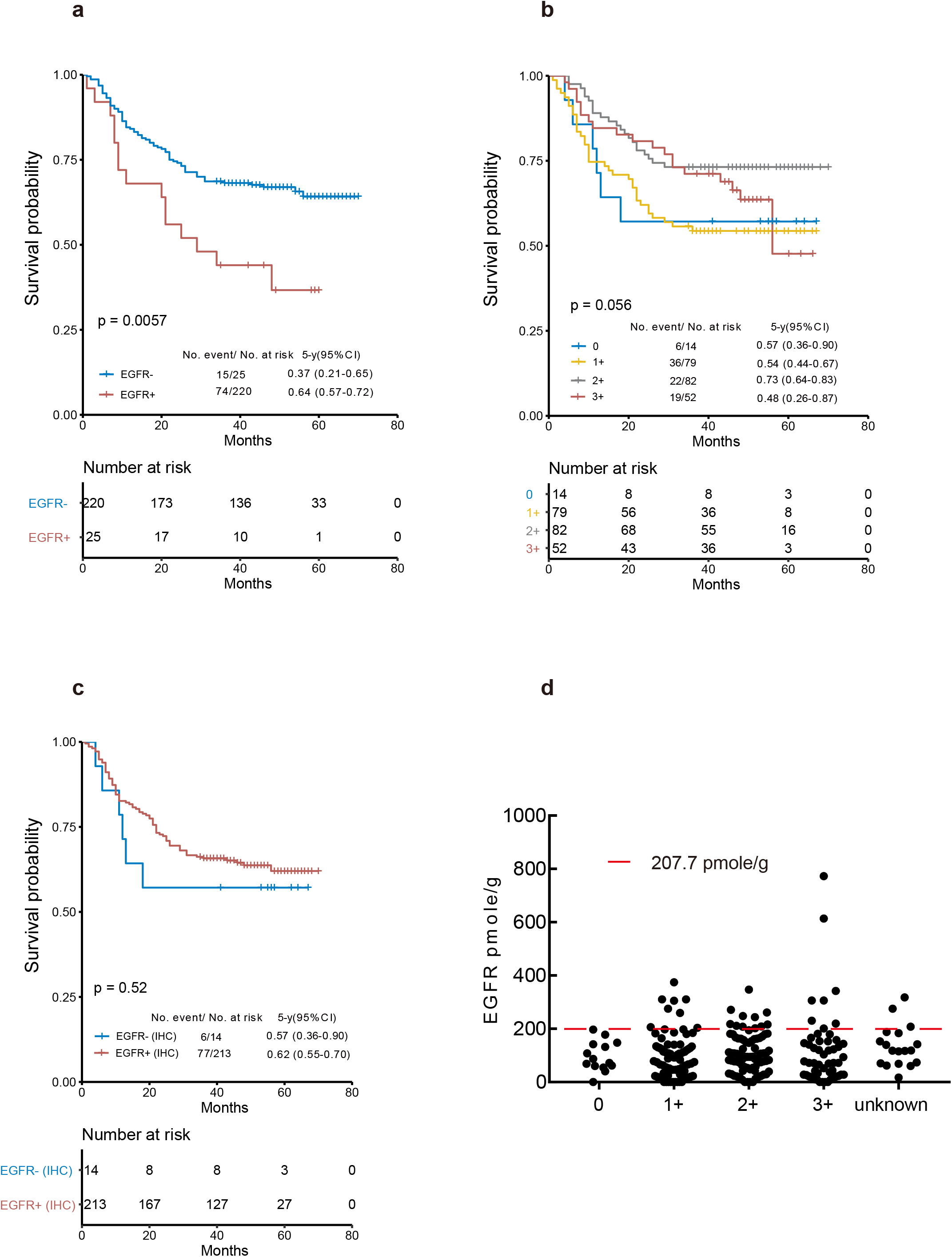
Kaplan-Meier OS analysis of gastric cancer patients based on their EGFR protein levels measured using QDB (**a**) or IHC methods (**b & c**). (**a**) Absolutely quantitated EGFR protein levels were used to stratify patients using a cutoff of 207.7 pmole/g into EGFR+ (n=25) and EGFR-groups (n=220) for Kaplan-Meier OS analysis, with 5y SP at 37% (95%CI: 21% to 65%) and 64% (95%CI: 57% to 72%) respectively. p=0.0057 from Log Rank test; (**b**) IHC scores of EGFR protein were used to stratify patients into 0 (n=14), 1+ (n=79), 2+ (n=82), and 3+ (n=52) groups for Kaplan-Meier OS analysis, with 5y SP at 57% (95%CI: 36% to 90%), 54% (95%CI: 44% to 67%), 73% (95%CI: 64% to 83%) and 48% (95% CI: 26% to 87%) respectively. p=0.056 from Log Rank test. (**c**) The patients were stratified using their IHC scores, with those of 0 as EGFR-, and those with IHC score ≥1+ as EGFR+ for Kaplan-Meier OS analysis, with 5y SP for EGFR-at 57% (95%CI: 36% to 90%), and for EGFR+ at 62% (95%CI: 55% to 70%). p=0.52 from Log Rank test. (d) The putative cutoff of 207.7 pmole/g was plotted on the distribution plot of EGFR protein levels sub-grouped by their IHC scores.

In Fig. **3b**, we also performed Kaplan-Meier analysis based on their IHC scores. Unexpectedly, opposite results were obtained using IHC scores as compared to those based on QDB measurements. The 2+ group patients were found with best 5y SP at 73% (95%CI: 64% to 83%), while those of 0, 1+ and 3+ were at 57% (95%CI: 36% to 90%), 54% (95%CI: 44% to 67%) and 48% (95%CI: 26% to 87%) respectively. The 3y SP of 3+ patients was highly similar to that of 2+ patients, with a sudden drop in next two years. When analyzed together, we achieved p=0.056 from Log Rank test.

In many clinical trials, all patients with IHC scores ≥1+ were considered EGFR+ patients. Therefore, the patients were stratified accordingly, and Kaplan-Meier OS analysis was performed in Fig. **3c**. We obtained p=0.52 from Log Rank test. Clearly, this stratification method was not effective for OS analysis of gastric patients. In Fig. **3d**, the putative cutoff used in 3a was plotted in the distribution plot of all gastric specimens sub-grouped by their IHC scores.

## Discussion

In this study, we presented the first practical method to measure EGFR protein levels in both gastric and Lung cancer tissues absolutely, objectively and quantitatively for routine clinical practice. The effectiveness of the QDB platform was demonstrated preliminarily by showing that EGFR was a negative prognostic factor for OS of gastric patients through statistical analysis; a conclusion cannot be repeated using IHC scores provided from the local hospital.

Our results also revealed the inadequacy of IHC method to provide a reliable assessment of EGFR levels among gastric patients. First, when measured EGFR levels absolutely and quantitatively, there were no statistical significant difference between any two groups of the 0/1+/2+/3+ groups categorized by IHC method using student t-test; second, the IHC scores of EGFR protein were not an independent prognostic factor when analyzed with Cox regression OS analysis (supplemental table 2); and third, when analyzed with Kaplan-Meier OS analysis, patients of 2+ group were found to be associated with the best 5y SP while those of 0,1+ and 3+ groups were highly similar among each other.

Perhaps the inadequacy of IHC method for proper stratification of patients is best illustrated in Fig. 3d. At least for prognostic purpose, while there were no EGFR+ specimens in 0 group, there were roughly equal numbers of EGFR+ within 1+, 2+ and 3+, with significantly more EGFR-specimens in all four IHC groups. The large number of EGFR-specimens within 1+, 2+ and 3+ groups should overwhelm any EGFR+-related events in clinical studies. The overall situation was not changed by setting the threshold at 2+ either. While we acknowledged these IHC scores from local hospital might lack the caliber equipped with the central laboratories in advanced countries, it nonetheless give us a glimpse of what may sabotage an otherwise successful clinical trial.

In fact, the limitation of IHC method has been well acknowledged, and efforts have been devoted to find a better alternative. Florescence in situ hybridization (FISH) technique is suggested to be a more consistent method to identify EGFR positive patients for Cetuximab or other anti-EGFR antibody drugs in several studies(5,12,23–25). Selective Reaction Monitoring-Mass spectrometry (SRM-MS) has also been used successfully to measure EGFR protein levels absolutely and quantitatively in NSCLC tissues(26). Additionally, the results from the SRM-MS study were comparable to our results. However, both these methods pose technical challenge for routine clinical practice. In addition, FISH method only detects changes at DNA level, and it is not a true quantitative method to allow defining an optimized cutoff through outcome analysis to guide patients for anti-EGFR therapies (12).

With the absolutely quantitated EGFR levels, we demonstrated clearly the prognostic role of EGFR protein in the OS of gastric cancer through both univariate and multivariate cox regression analysis. We also proposed a putative cutoff of 207.7 pmole/g through outcome analysis for prognosis of gastric cancer patients. Caution interpretation of this putative cutoff is warranted, as its validity remains to be confirmed in an independent cohort of patients.

It remains to be seen if the above-mentioned cutoff was the same as the one we were seeking to guide patient for anti-EGFR therapies. Clearly, it can only be answered through outcome analysis [progression free survival (PFS), OS or Overall Response Rate (ORR)] of the patients receiving anti-EGFR therapies in retrospective and/or prospective clinical trials in the near future.

Nonetheless, we showed clearly in this study that an optimized cutoff can be identified through outcome analysis using absolutely quantitated EGFR levels. Thus, the threshold to stratify patients will no longer be set arbitrarily, but rather objectively through outcome analysis (PFS, OS, or ORR) of patients receiving anti-EGFR therapies. The absolute nature of the QDB results can also enable seamless integration of multiple clinical studies to expand the scale of the study, and the derived optimized cutoff may be accordingly fine-tuned to identify more precisely the patient population who may truly benefit from anti-EGFR therapies in the near future.

We noticed the range of EGFR protein expression among gastric patients was much narrower than that of Lung cancer patients. The expression levels of EGFR in both tissues were also much lower than those of Her2 protein in breast cancer tissues. In several studies including our own, Her2 distribution was from undetectable level to 31.31 nmole/g, with the cutoff to match the results from IHC/FISH at 0.261 nmole/g when measured with an anti-Her2 (EP3) antibody (21). In contrast, the distribution of EGFR was from 0 to 0.773 nmole/g (n=246) in gastric cancer specimens, and from 0 to 2.695 nmole/g (n=81) in Lung cancer specimens. We speculate that is why the success of IHC-based Her2 assessment cannot be duplicated with EGFR protein. The narrow range of EGFR protein levels in either gastric or lung cancer tissues may pose a significant technical challenge to IHC-based EGFR assessment in current clinical practice.

In conclusion, a QDB-based immunoassay was introduced to measure EGFR protein levels absolutely, quantitatively and objectively in both gastric and Lung cancer specimens. Validation and adoption of this assay should help define EGFR protein as a predictive biomarker to accelerate clinical trials aiming to extend anti-EGFR therapies, including Cetuximab, Panitumumab, and necitumumab, in gastric and lung cancers.

## Data Availability

Data are available from the correspondent author upon reasonable written request.

## Acknowledgement

The authors wish to thank Ms. Wenfeng Zhang, Ms. Yan Lv & Ms. Yunyun Zhang for their excellent technical supports.

## Author contributions

LX & BS provided clinical specimens and matching clinicopathological information, and supervised all the clinical studies; FT performed all the assays and performed data analysis; MY & FT contributed to data interpretation and edited the manuscript; JZ designed & supervised the overall study and drafted the manuscript.

## Data Availability

Data are available from the correspondent author upon reasonable written request.

## Reference

1. Adashek JJ, Arroyo-Martinez Y, Menta AK, Kurzrock R, Kato S. Therapeutic Implications of Epidermal Growth Factor Receptor (EGFR) in the Treatment of Metastatic Gastric/GEJ Cancer. Front Oncol [Internet]. Frontiers; 2020 [cited 2021 Mar 25];10. Available from: https://www.frontiersin.org/articles/10.3389/fonc.2020.01312/full

2. Chae YK, Arya A, Chiec L, Shah H, Rosenberg A, Patel S, et al. Challenges and future of biomarker tests in the era of precision oncology: Can we rely on immunohistochemistry (IHC) or fluorescence in situ hybridization (FISH) to select the optimal patients for matched therapy? Oncotarget. 2017;8:100863–98.

3. Ai X, Guo X, Wang J, Stancu AL, Joslin PMN, Zhang D, et al. Targeted therapies for advanced non-small cell lung cancer. Oncotarget. 2018;9:37589–607.

4. Díaz-Serrano A, Sánchez-Torre A, Paz-Ares L. Necitumumab for the treatment of advanced non-small-cell lung cancer. Future Oncol. 2019;15:705–16.

5. Han S-W, Oh D-Y, Im S-A, Park SR, Lee K-W, Song HS, et al. Phase II study and biomarker analysis of cetuximab combined with modified FOLFOX6 in advanced gastric cancer. Br J Cancer. 2009;100:298–304.

6. Licitra L, Störkel S, Kerr KM, Van Cutsem E, Pirker R, Hirsch FR, et al. Predictive value of epidermal growth factor receptor expression for first-line chemotherapy plus cetuximab in patients with head and neck and colorectal cancer: analysis of data from the EXTREME and CRYSTAL studies. Eur J Cancer. 2013;49:1161–8.

7. Lordick F, Kang Y-K, Chung H-C, Salman P, Oh SC, Bodoky G, et al. Capecitabine and cisplatin with or without cetuximab for patients with previously untreated advanced gastric cancer (EXPAND): a randomised, open-label phase 3 trial. The Lancet Oncology. 2013;14:490–9.

8. Pirker R, Pereira JR, von Pawel J, Krzakowski M, Ramlau R, Park K, et al. EGFR expression as a predictor of survival for first-line chemotherapy plus cetuximab in patients with advanced non-small-cell lung cancer: analysis of data from the phase 3 FLEX study. The Lancet Oncology. 2012;13:33–42.

9. Thatcher N, Hirsch FR, Luft AV, Szczesna A, Ciuleanu TE, Dediu M, et al. Necitumumab plus gemcitabine and cisplatin versus gemcitabine and cisplatin alone as first-line therapy in patients with stage IV squamous non-small-cell lung cancer (SQUIRE): an open-label, randomised, controlled phase 3 trial. The Lancet Oncology. Elsevier; 2015;16:763–74.

10. Burtness B, Goldwasser MA, Flood W, Mattar B, Forastiere AA. Phase III Randomized Trial of Cisplatin Plus Placebo Compared With Cisplatin Plus Cetuximab in Metastatic/Recurrent Head and Neck Cancer: An Eastern Cooperative Oncology Group Study. JCO. Wolters Kluwer; 2005;23:8646–54.

11. Lordick F, Luber B, Lorenzen S, Hegewisch-Becker S, Folprecht G, Wöll E, et al. Cetuximab plus oxaliplatin/leucovorin/5-fluorouracil in first-line metastatic gastric cancer: a phase II study of the Arbeitsgemeinschaft Internistische Onkologie (AIO). Br J Cancer. 2010;102:500–5.

12. Luber B, Deplazes J, Keller G, Walch A, Rauser S, Eichmann M, et al. Biomarker analysis of cetuximab plus oxaliplatin/leucovorin/5-fluorouracil in first-line metastatic gastric and oesophago-gastric junction cancer: results from a phase II trial of the Arbeitsgemeinschaft Internistische Onkologie (AIO). BMC Cancer. 2011;11:509.

13. Tol J, Dijkstra JR, Klomp M, Teerenstra S, Dommerholt M, Vink-Börger ME, et al. Markers for EGFR pathway activation as predictor of outcome in metastatic colorectal cancer patients treated with or without cetuximab. Eur J Cancer. 2010;46:1997–2009.

14. Paz-Ares L, Mezger J, Ciuleanu TE, Fischer JR, von Pawel J, Provencio M, et al. Necitumumab plus pemetrexed and cisplatin as first-line therapy in patients with stage IV non-squamous non-small-cell lung cancer (INSPIRE): an open-label, randomised, controlled phase 3 study. The Lancet Oncology. 2015;16:328–37.

15. Apicella M, Corso S, Giordano S. Targeted therapies for gastric cancer: failures and hopes from clinical trials. Oncotarget. 2017;8:57654–69.

16. Li K, Li J. Current Molecular Targeted Therapy in Advanced Gastric Cancer: A Comprehensive Review of Therapeutic Mechanism, Clinical Trials, and Practical Application. Gastroenterology Research and Practice. Hindawi; 2016;2016:e4105615.

17. Lynch TJ, Patel T, Dreisbach L, McCleod M, Heim WJ, Hermann RC, et al. Cetuximab and First-Line Taxane/Carboplatin Chemotherapy in Advanced Non– Small-Cell Lung Cancer: Results of the Randomized Multicenter Phase III Trial BMS099. Journal of Clinical Oncology [Internet]. American Society of Clinical Oncology; 2010 [cited 2021 Mar 31]; Available from: https://ascopubs.org/doi/pdf/10.1200/JCO.2009.21.9618

18. Gown AM. Diagnostic Immunohistochemistry: What Can Go Wrong and How to Prevent It. Archives of Pathology & Laboratory Medicine. 2016;140:893–8.

19. Tian G, Tang F, Yang C, Zhang W, Bergquist J, Wang B, et al. Quantitative dot blot analysis (QDB), a versatile high throughput immunoblot method. Oncotarget. 2017;8:58553–62.

20. Zhang W, Yu G, Zhang Y, Tang F, Lv J, Tian G, et al. Quantitative Dot Blot (QDB) as a universal platform for absolute quantification of tissue biomarkers. Analytical Biochemistry. 2019;576:42–7.

21. Yu G, Zhang W, Zhang Y, Lv J, Wu S, Sui X, et al. Developing a routine lab test for absolute quantification of HER2 in FFPE breast cancer tissues using Quantitative Dot Blot (QDB) method. Sci Rep. 2020;10:12502.

22. Hao J, Lv Y, Zou J, Zhang Y, Xie S, Jing L, et al. Improving prognosis of surrogate assay for breast cancer patients by absolute quantitation of Ki67 protein levels using Quantitative Dot Blot (QDB) method. medRxiv. Cold Spring Harbor Laboratory Press; 2020;2020.03.11.20034439.

23. Genova C, Socinski MA, Hozak RR, Mi G, Kurek R, Shahidi J, et al. EGFR Gene Copy Number by FISH May Predict Outcome of Necitumumab in Squamous Lung Carcinomas: Analysis from the SQUIRE Study. J Thorac Oncol. 2018;13:228–36.

24. Hirsch FR, Herbst RS, Olsen C, Chansky K, Crowley J, Kelly K, et al. Increased EGFR gene copy number detected by fluorescent in situ hybridization predicts outcome in non-small-cell lung cancer patients treated with cetuximab and chemotherapy. J Clin Oncol. 2008;26:3351–7.

25. Herbst RS, Redman MW, Kim ES, Semrad TJ, Bazhenova L, Masters G, et al. Cetuximab plus carboplatin and paclitaxel with or without bevacizumab versus carboplatin and paclitaxel with or without bevacizumab in advanced NSCLC (SWOG S0819): a randomised, phase 3 study. Lancet Oncol. 2018;19:101–14.

26. Hembrough T, Thyparambil S, Liao W-L, Darfler MM, Abdo J, Bengali KM, et al. Selected Reaction Monitoring (SRM) Analysis of Epidermal Growth Factor Receptor (EGFR) in Formalin Fixed Tumor Tissue. Clin Proteomics. 2012;9:5.

