## Supplemental table 1, 2 and Supplemental Fig. 1 for "Objective quantitation of EGFR protein levels using Quantitative Dot Blot (QDB) method for prognosis of gastric cancer patients"

**Supplemental table 1：Clinicopathological characteristics of lung cancer specimens.**

|  | Cases（n=81） | |
| --- | --- | --- |
| **Characteristics** | n | % |
| **Age** |  |  |
| <60 | 20 | 24.69 |
| ≥60 | 50 | 61.73 |
| unknown | 11 | 13.58 |
| **Gender** |  |  |
| Male | 45 | 55.56 |
| Female | 25 | 30.86 |
| unknown | 11 | 13.58 |
| **Histopathological type** |  |  |
| Small cell carcinoma | 1 | 1.23 |
| adenocarcinoma | 45 | 55.56 |
| Squamous cell carcinomas | 19 | 23.46 |
| Large cell carcinoma | 2 | 2.47 |
| other | 2 | 2.47 |
| unknown | 12 | 14.81 |
| **Tumor size(cm)** |  |  |
| ≤3 | 9 | 11.11 |
| 3~5 | 5 | 6.17 |
| >5 | 4 | 4.94 |
| unknown | 63 | 77.78 |
| **Lymph node metastasis** |  |  |
| No metastasis | 9 | 11.11 |
| Metastasis | 5 | 6.17 |
| unknown | 67 | 82.72 |

**Supplemental table 2: Univariate and multivariate cox regression OS analysis of gastric cancer patients using IHC scores of EGFR protein.**

|  | Univariate | | |  | Multivariate | | |
| --- | --- | --- | --- | --- | --- | --- | --- |
| Variable | HR | 95%CI | P.value |  | HR | 95%CI | P.value |
| Gender | 1.44 | 0.86-2.41 | 0.1677 |  | 1.81 | 0.98-3.34 | 0.0577 |
| Age | 1.11 | 0.72-1.73 | 0.6339 |  | 0.97 | 0.58-1.63 | 0.9153 |
| Tumor size | 1.74 | 1.13-2.68 | 0.0118 |  | 1.2 | 0.72-1.98 | 0.4832 |
| Lauren's classification | 1.39 | 1.04-1.85 | 0.0249 |  | 1.09 | 0.72-1.63 | 0.692 |
| Histologic grade | 1.8 | 0.79-4.12 | 0.1647 |  | 0.52 | 0.18-1.46 | 0.2135 |
| T-stage | 1.96 | 1.51-2.53 | 0 |  | 1.35 | 0.93-1.97 | 0.1152 |
| LNsN | 1.88 | 1.55-2.27 | 0 |  | 1.43 | 1.10-1.86 | 0.0077 |
| Vascular cancer embolus | 3.45 | 2.12-5.61 | 0 |  | 1.73 | 0.94-3.17 | 0.0794 |
| Nerve invasion | 3.03 | 1.95-4.72 | 0 |  | 1.21 | 0.67-2.20 | 0.5339 |
| EGFR_IHC_ | 0.76 | 0.33-1.75 | 0.5244 |  | 1.02 | 0.43-2.43 | 0.9571 |

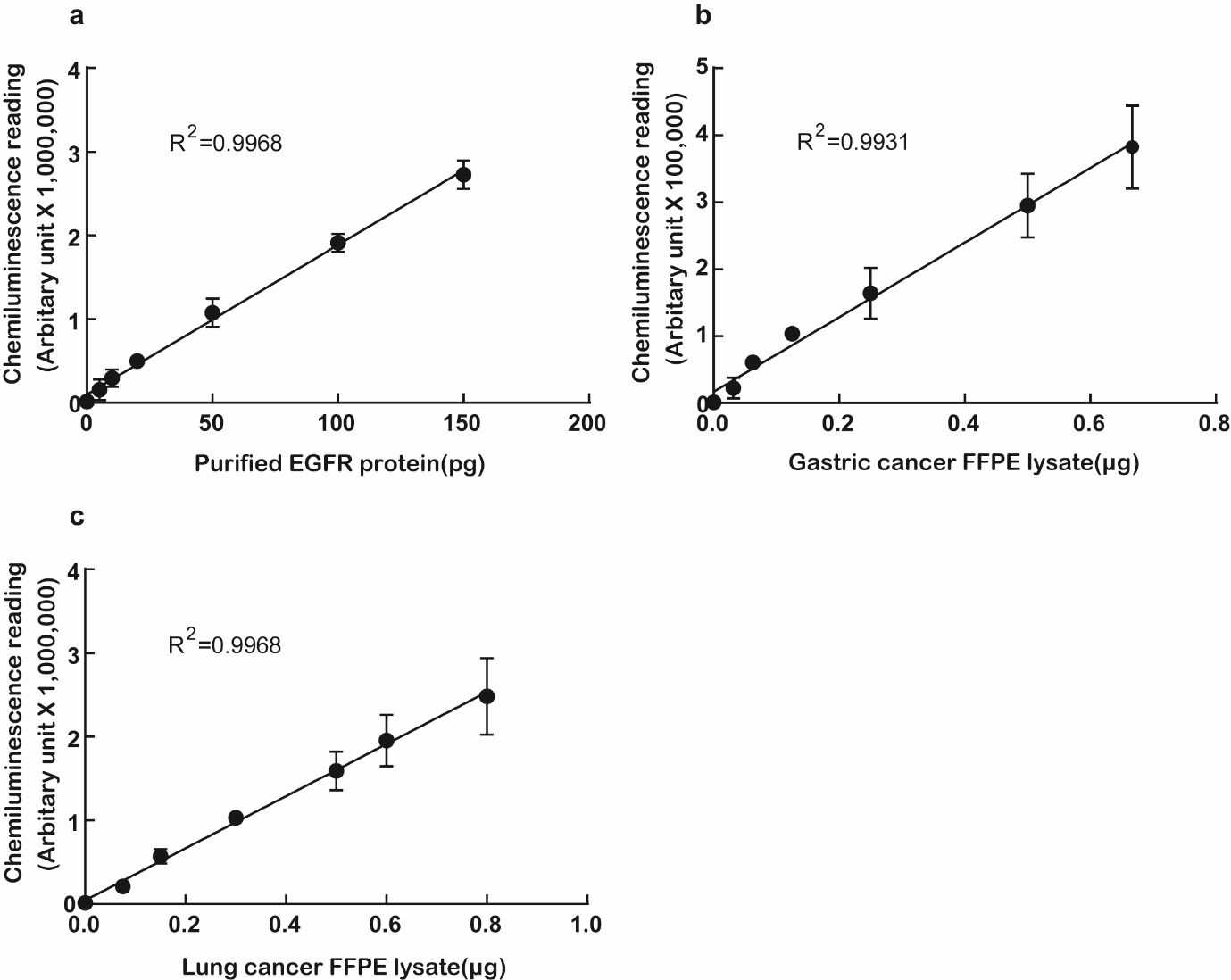

**Supplemental figure 1: Defining the linear range of QDB measurements with anti-EGFR antibody, EP22.** (**a**) Defining the linear range of QDB method for analysis of recombinant human EGFR protein. The purified EGFR protein was serially diluted with 0.5 µg/µl IgG-free BSA solution. The diluted solution was used for QDB analysis in triplicate. (**b, c**) Defining the linear range of QDB method for analysis of EGFR protein levels in gastric and lung cancer FFPE tissue lysates. Five gastric or lung cancer FFPE tissue lysates of moderate expression of EGFR protein were pooled together. The pooled lysate was serially diluted as indicated in the figure, supplemented with 0.5 µg/µl IgG-free BSA solution to ensure equal loading of the samples. The total samples were applied onto the QDB plate in triplicate for QDB analysis. The results were expressed as average ± SD (standard deviation).
